# Suspected rabies exposure among animal-bite human cases in Busia district, Uganda: Prevalence, associated factors and delayed post-exposure care-seeking. A cross-sectional study

**DOI:** 10.64898/2026.05.29.26354408

**Authors:** David Wagaba, Immaculate Nabukenya, Jimmy Kizza, Happy Unith, Angel Kanyange, Christine Turyahabwe, Henry Kibuuka, Derrick Mugisha, Shamim Nabidda, Simon Peter Ogola, Lois Keren Kisakye, Joan N Kalyango

## Abstract

**Background:** Rabies is a zoonotic neglected public health problem associated with animal bites, especially domestic carnivores claiming 59,000 deaths annually predominantly in developing countries of Africa and Asia. There is a high risk of exposure among rural communities endemic with animal rabies where adoption of prevention strategies is minimal. This study determined the prevalence of suspected rabies exposure, associated factors, and delayed post-exposure care-seeking among animal-bite human cases in Busia district, Uganda.

**Methods:** This was a cross-sectional study that involved 332 consecutively sampled animal bite human cases that occurred within the period 2023 to 2024. Data for the bite cases from records were collected using a data abstraction tool. In addition, interviewer-administered semi-structured questionnaires were used to collect data on sociodemographic, animal-related and environmental characteristics. Approximate bite locations were collected using Global Positioning System (GPS) coordinates via Kobo collect. Analysis was carried out in STATA 17 using mixed effects modified Poisson regression for factors associated with suspected rabies exposure.

**Results:** The median age of the bite cases was 18 (IQR: 9-36) with the male gender predominantly affected. The prevalence of suspected rabies exposure was 53.6% (95% Confidence interval - CI: 46.8-60.3). Factors associated were urban versus (vs) rural residence (adjusted prevalence ratio-aPR: 1.04, 95%CI: 1.00-1.08), being bitten by a stray animal (aPR: 1.28, 95% CI: 1.22-1.35) and wild animal (aPR: 1.22, 95% CI: 1.14-1.30) vs domestic animal, vaccination status of the biting animal i.e. vaccinated vs unvaccinated (aPR: 0.76, 95% CI: 0.69-0.85), provoked vs unprovoked bites (aPR: 0.82, 95% CI: 0.79-0.86), and distance to nearest river (≥5km) vs <5km (aPR: 0.93, 95% CI: 0.87-0.99). The prevalence of delayed post-exposure seeking was 23.0% (95% CI: 16.5-31.1) among the suspected rabies exposures.

**Conclusion:** The study reveals a high prevalence of suspected rabies exposure. Factors associated are multidimensional i.e. are of human, animal and environmental origin. The one health paradigm should be emphasized during routine surveillance of rabies-related cases. The study observed that 1 in 5 bite cases delayed to seek care post bite exposure. We recommend collaborations between sectors, routine vaccination and awareness campaigns, and monitoring of wild carnivore populations and environmental dynamics in rabies-related surveillance.

**Author Summary:** Rabies continues to be a neglected public health problem in sub-Saharan Africa. Vaccination and treatment following an animal bite is important to prevent clinical disease. Such treatment is costly in developing countries necessitating the use of risk assessment to identify potential suspected cases. We investigated the occurrence of suspected rabies exposures, associated factors and delayed care seeking around bite incidents in a district in Uganda with the highest animal rabies reports. Our study revealed that more than half of the bite cases were suspected to be exposed. Factors associated were of human, animal and environmental related origin. These were urban residence, stray and wild animal bites, provocation status of the bite, type of animal and distance to the nearest river. At least 1 in 5 individuals with suspected rabies exposures delayed to seek for care following the bite incident. Our findings highlight the need for collaboration between stakeholders in the human, animal and environmental sectors to contribute to the eradication of rabies and related instances by 2030.

## Introduction

Rabies is a zoonotic viral disease of public health importance caused by a lyssavirus of the virus family *Rhabdoviridae* (1) transmitted through bites by rabid animals most especially dogs. Transmission commonly occurs through direct contact of saliva from infected animals with fresh wounds or mucosal surfaces. Transmission via transdermal scratches is also known to occur. Case fatality is almost 100% by the time clinical manifestation occurs (2).

Globally, 4.5 million people are bitten by dogs (3) with about 59,000 rabies mortalities occurring annually, registered predominantly in the developing countries of Africa and Asia (1). Rabies associated mortalities in Africa account for about 40% (i.e. 21,000-25,000) of the global rabies mortality estimates (4). The persistence of rabies in Africa has been attributed to a lack of accurate epidemiological data, negligence by populations and decision-makers, lack of awareness of rabies risk, as well as geographical and financial barriers to post-exposure prophylaxis (PEP) access (5). Between 2015 and 2020, Uganda recorded 14,865 dog bites and 36 rabies deaths annually (6). These patterns are postulated to be maintained by high poverty and low literacy levels in rural areas where majority of the outbreaks occur. This results in less adoption of preventive strategies such as rabies vaccination and breeding control leading to an increase in the number of unvaccinated dogs (7). Poor animal welfare encourages straying away of domesticated pets to environmental habitats in search of food and mates. As a result, domesticated pets interact with wildlife, enhancing the risk of acquiring rabies;relaying the infection to human populations (8). Additionally, it is reported that only 10% of domesticated dogs are vaccinated against rabies in Uganda compared to the 70% recommendation by the World Health Organization (WHO) required for herd immunity (6). This results in less protection among the domesticated pets with a high potential of becoming rabid once infected. This places humans in endemic settings like Busia district at risk. Busia district was reported to have had the highest animal rabies prevalence of 14% among all districts in Uganda (9).

Following exposure, timely administration of rabies PEP with good efficacy is critical for preventing clinical rabies (10). Economic losses due to rabies have been estimated to amount to about 8.6 billion US dollars (2). These economic losses arise from deaths of infected animals, the cost of PEP, income loss for animal bite victims and other related costs. In light of the high PEP costs especially in low-income countries, the WHO developed risk assessment guidelines to prioritize PEP administration for individuals with suspected rabies exposure. This entailed prioritization of category II (minor scratches, abrasions without bleeding, or nibbling of uncovered skin) and III (transdermal bites or scratches, licks on broken skin, or mucous membrane contamination) bite exposures in addition to information about the animal’s disposition with regard to rabies (1). This aimed to limit unwarranted recommendation of rabies PEP that could result in over-prescription of these biologics (11). This is also supported by the fact that even in endemic settings, not every bite exposure is necessarily linked to a rabid animal (1).

Rabies occurrence has been associated with factors such as age group, gender, socio-economic status, and level of education among others (1). Spatial and temporal characteristics have also been reported to influence rabies occurrence, however, studies exhibiting these associations are limited. Some of these factors include environmental temperature, rainfall/precipitation, elevation, presence of nearby waterbodies, and population density (12).

Rural communities are highly affected by rabies outbreaks because of limited access to health care, low socio-economic status, and a threat to their livelihoods due to attacks on livestock (13). Domestic-wild animal interactions maintain the endemic status of the disease in such areas. The effectiveness of proposed preventive strategies is hindered by long distances to health facilities, low public awareness and scarce resources supplied to these settings leading to undesirable consequences (14).

The WHO established a strategic goal to end rabies deaths by 2030. Suggested objectives included reducing rabies risk, providing guidance and data, and harnessing multi-stakeholder engagement. It was also emphasized that achieving this goal required the use of the One health approach i.e. collaboration between human, animal and environmental health sectors (15). Rabies risk management has been shown to require, at a minimum, collaboration between the human and animal health sectors; a practice present in Busia district, Uganda. The Uganda One health Platform established the need to establish the vulnerability within endemic communities and map the risk (rabies exposure) in relation to the environment through multidisciplinary research (16). This follows the observation from previous research that rabies occurrence in the country is associated with environmental factors such as weather changes, geographical location and proximity to wildlife habitats (8, 17)

To facilitate appropriate planning, comprehensive epidemiological information on suspected rabies exposure in such areas using the One Health perspective is recommended. Therefore, this study aimed to determine the prevalence of suspected rabies exposure and associated factors among animal-bite human cases in Busia district. Additionally, this study described the prevalence of delayed post-exposure care-seeking among those with suspected rabies exposure.

## Materials and Methods

### Study design and setting

This was a cross-sectional study which involved review of 332 human animal-bite records at Busia district veterinary office registered from 1^st^ January 2023 to 31^st^ December 2024. Busia district is located in the south-eastern part of Uganda, north of Lake Victoria and west of the Republic of Kenya approximately 196 kilometres (km) from Kampala the capital city of Uganda. It is bordered by Tororo district to the north, Busia County, Kenya to the east, Namayingo district to the south-west, and Bugiri district to the west. The district lies approximately between longitudes 33.05’ East and 34.01’ East, and latitude 00.10’North and 00.35’ North and it covers a total surface area of 743 km^2^. Busia is a one-county district (Samia-Bugwe County) and has one municipal council (Busia Municipal Council), 14 sub-counties, two divisions (Eastern and Western), 63 parishes and 534 (509 rural & 25 urban) villages. Busia district has one general hospital (Masafu Hospital, one health centre IV (Busia Health center IV), eleven health centre IIIs.

### Integrated bite case management in Busia district

Following an animal bite, victims may visit the nearest health centre or interface with a trained community health worker (CHW) for first aid care. Information about the animal bite incident is then communicated to the district veterinary office (DVO) for rabies risk assessment on the basis of the animal disposition. Thereafter, the animal bite victim has to visit the DVO for information as to whether they will be referred for PEP rabies vaccination (i.e. suspect for rabies exposure) or reception of a tetanus shot. This information is captured on a referral form which is issued by the DVO. If a victim is deemed suspect for rabies exposure, they are referred to a health facility for PEP.

### Study Population

This comprised of animal-bite human cases that had records available at the DVO reported within the period from 1^st^ January 2023 to 31^st^ December 2024. We included only residents of Busia district with active telephone contacts and consented to the study. Animal bite case records for people who were not reachable on phone or were too ill to respond were excluded.

### Sample size and sampling procedure

The sample size for the prevalence of suspected human rabies exposure, was calculated using the formula for a single proportion (18) with a design effect of 2 to account for anticipated clustering within the data. The computation was based on a 95% level of confidence, 5% tolerable sampling error and estimated prevalence of 10.47% from a previous study (19). Based on the computation, the required sample size was approximately 289 animal bite records. The sample size for factors associated was computed using the formula for comparing two proportions (18). It assumed a 5% level of significance, 80% power of the study using proportion estimates from a previous study (20). These included proportion of subjects bitten by domestic pets; 0.442 (q1), proportion of subjects bitten by free-roaming/stray animal; 0.558 (q2), proportion of subjects bitten by domestic pets with rabies exposure; 0.0594 (p1) and proportion of subjects bitten by free-roaming/stray animal with rabies exposure; 0.2706 (p2). Based on the computation, the required sample size was approximately 196 records. Sample size estimation was not considered for delayed post-exposure care-seeking as it was a descriptive objective secondary to those with suspected rabies exposure. Therefore, the larger sample size of 289 was chosen and adjusted for a 10% missing data rate and a 5% contact failure rate, yielding a final sample size of 332 animal bite records. A proportionate stratified sampling procedure was employed, with strata based on months of the year to ensure representative coverage. Records were organized by year, then by month, and proportions were allocated accordingly. Within each month, records were further organized by subcounty, and consecutive sampling was applied to select the required records.

### Study Variables

Dependent variables included Suspected rabies exposure defined as animal-bite human case that were referred for Post Exposure Prophylaxis rabies vaccination following risk assessment by a qualified veterinarian following bite exposure as per the Uganda Clinical Guidelines 2023 (21). Some of the aspects considered include;

#### Nature of Contact

Exposure is classified into three bite categories as per WHO guidelines: Category I (touching or feeding animals, licks on intact skin; no exposure), Category II (minor scratches or abrasions without bleeding, or nibbling of uncovered skin; minor exposure), and Category III (single or multiple transdermal bites or scratches, licks on broken skin, or contamination of mucous membranes with saliva; severe exposure).

#### Animal Status

The implicated animal is categorized based on its vaccination history, availability for a 10-day veterinary observation period, and clinical presentations of rabies (e.g., unprovoked aggression, hyper-salivation, or paralysis). Bites involving untraceable strays, wild carnivores, or symptomatically abnormal animals elevate the level of suspicion.

#### Anatomical Site

Injuries are categorized based on proximity to the central nervous system and nerve density, with wounds to the head, neck, face, and hands classified as high-priority due to their potential for a shortened incubation period.

The other dependent variable was delayed post-exposure care seeking defined as an interval exceeding 48 hours between dates of bite exposure and PEP referral (1, 22)

Independent variables included **sociodemographic characteristics:** age, sex, family size, highest level of education, occupation of household head, family size, marital status of household head, residence.

#### Animal-related factors

pet ownership, pet vaccination status, type of animal, presence of wild carnivore in vicinity, breeding control, livestock ownership, number of bites, provocation status, time of day, and bite location from home

#### Ecological/Environmental factors

elevation, distance to main rivers, temperature, rainfall amount, normalised difference vegetation index (NDVI), presence of forest nearby, nearby road access.

### Data collection procedure

Data was collected by the principal investigator with support of three (3) trained research assistants comprised of two para-veterinarians and a community health worker. The first phase involved abstraction of contact information from the referral forms to reach out to the potential study participants and obtain approximate location information after verbal consent.

The second phase involved reaching out to the participants’ homes and a semi-structured questionnaire in Kobo collect was administered through interviews in either English, Samia, Swahili or Luganda according to the participants’ preference. The corresponding identification numbers (IDs) were maintained on the questionnaires for each participant interviewed to ensure consistency within the data. Global Positioning System (GPS) coordinates for the approximate bite locations were taken using the Kobo collect tool following information provided by the participant. If the bite incident did not occur around the participants residence, details on the bite location were inquired and visited to capture the corresponding GPS coordinate.

QGIS version 3.40.3 was used to collect geographical/environmental variables with reference to the coordinate points. Monthly rainfall data was collected from google climate engine using the CHIRPS rainfall raster files at a resolution of 4.8km. Monthly temperature was collected as land surface temperature using the google climate engine using raster files with resolution of 30m. Zonal statistics were used to obtain rainfall and temperature estimates within 5km radius. This was also applied to obtain NDVI estimates within a 1km radius. Euclidean distances for distances to nearby physical features were also calculated in QGIS using nearest distance algorithm.

### Data management and analysis

Dataset preparation and cleaning was carried out in R software. The cleaned data was then exported to STATA 17 MP software for analysis. Continuous variables were described using medians and interquartile ranges (IQR). Frequencies and percentages were used to describe the categorical independent variables. Graphics such as bar graphs and maps were used to describe the study population characteristics. Due to the anticipation of clustering within the data at subcounty and month levels, analysis was carried out using survey data methods. Prevalence estimates were obtained by computing the proportions of the cases identified with suspected rabies exposure or delay post-exposure care-seeking. For factors associated with suspected rabies exposure, multi-level mixed effects modified Poisson regression (Poisson regression while using robust standard errors) was used to estimate the prevalence ratios for each of the independent variables. Independent variables with p-values less than 0.2 at bivariate analysis were selected for multivariable analysis. In multivariable analysis, interaction was assessed by carrying out the likelihood ratio (chunk) test. Interaction terms were made between significant variables after running a manual stepwise analysis for the variables selected at bivariate analysis. No statistical interaction was observed in the data since the chunk test was insignificant. Using manual stepwise estimation methods, independent variables significantly associated with suspected rabies exposure (p-value <0.05) were determined. Confounding on these significant independent variables was checked using the variables dropped during the stepwise estimation starting with the last dropped. An independent variable was considered a confounder if it resulted in a more than ten percent change (>10%) in the prevalence ratio (PR) for any of the significant independent variables. However, no variables were identified as confounders in the data. Therefore, the final model comprised of the significant independent variables.

## Results

Figure 1 below shows the study profile of data collected among the animal-bite human cases in Busia District reported between 1^st^ January 2023 and 31^st^ December 2024 at the DVO. Five hundred and twenty-five bite case records were accessed. Of these 20 did not have date and/or contact information and therefore did not meet the eligibility criteria. The remaining 505 bite case records were subjected to consecutive sampling to obtain the 332 records for study. Only one participant was excluded because they were ill.

**Figure 1:**
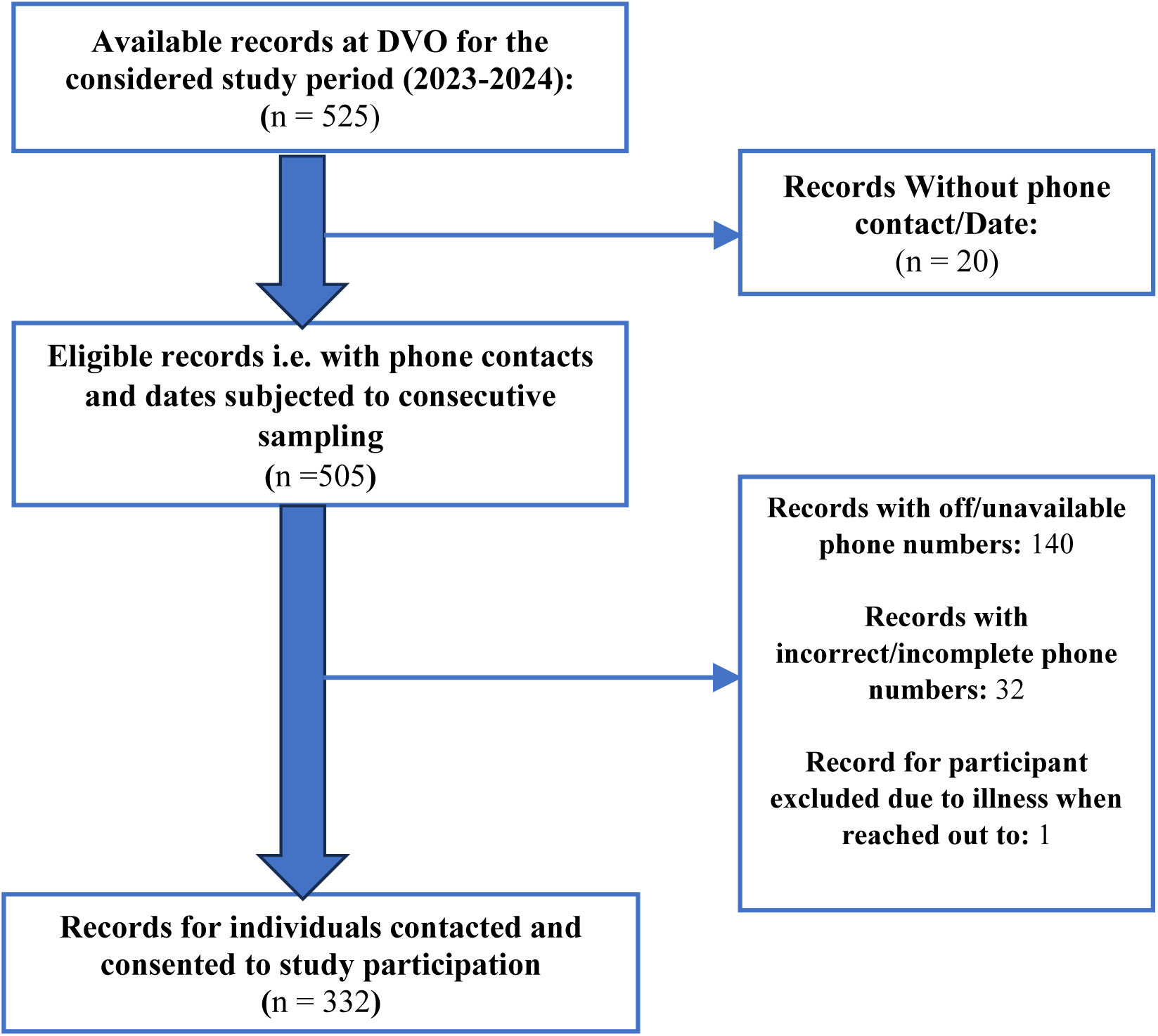
Study Profile

### Sociodemographic characteristics

The median age of the participants was 18 years (IQR: 9-36). More than half of the participants had household heads aged between 36 to 59 years (55.7%, n=185). Majority of the participants were of the male (61.4%, n=204), had attained primary level education (57.8%, n=192), and resided in rural areas (88.9%, n=295). Households had a median family size of 6 (IQR: 4-9) with the majority headed by married individuals (87.3%, n=290). The socio-demographic characteristics are summarised in **Error! Reference source not found.** below.

### Animal-related and environmental characteristics

Majority of the bites were inflicted by domesticated animals (67.8%, n=225) and were single bite exposures (95.2%, n=316). More than half of the bite incidents were unprovoked (53.0%, n=176), with 17.5% (n=58) of the biting animals reported to have been vaccinated against rabies. Approximately one-third of the participants reported pet ownership (28.3%, n=94) of which more than half reported having vaccinated them against rabies. The median elevation for the bite case locations was 1.19km (IQR: 1.17-1.21). The mean land surface temperatures had a median of 33.5°C (IQR: 31.6-35.2). A large portion of the bite cases occurred far away (>=5km) from the forest (88.0%, n=292) and near a river (72.6%, n=241). Most bite cases occurred within a 5km distance from the nearest main road (86.7%, n=288). Mean NDVI values within 1km radius of the bite case location had a median of 0.31 (IQR: 0.27-0.34). More than half of the bite cases occurred within the dry season (51.8%, n=172). **Error! Reference source not found.** below summarises the animal-related and environmental characteristics.

The majority of the bite cases reported were inflicted by dogs as shown in Figure 2 below.

**Figure 2:**
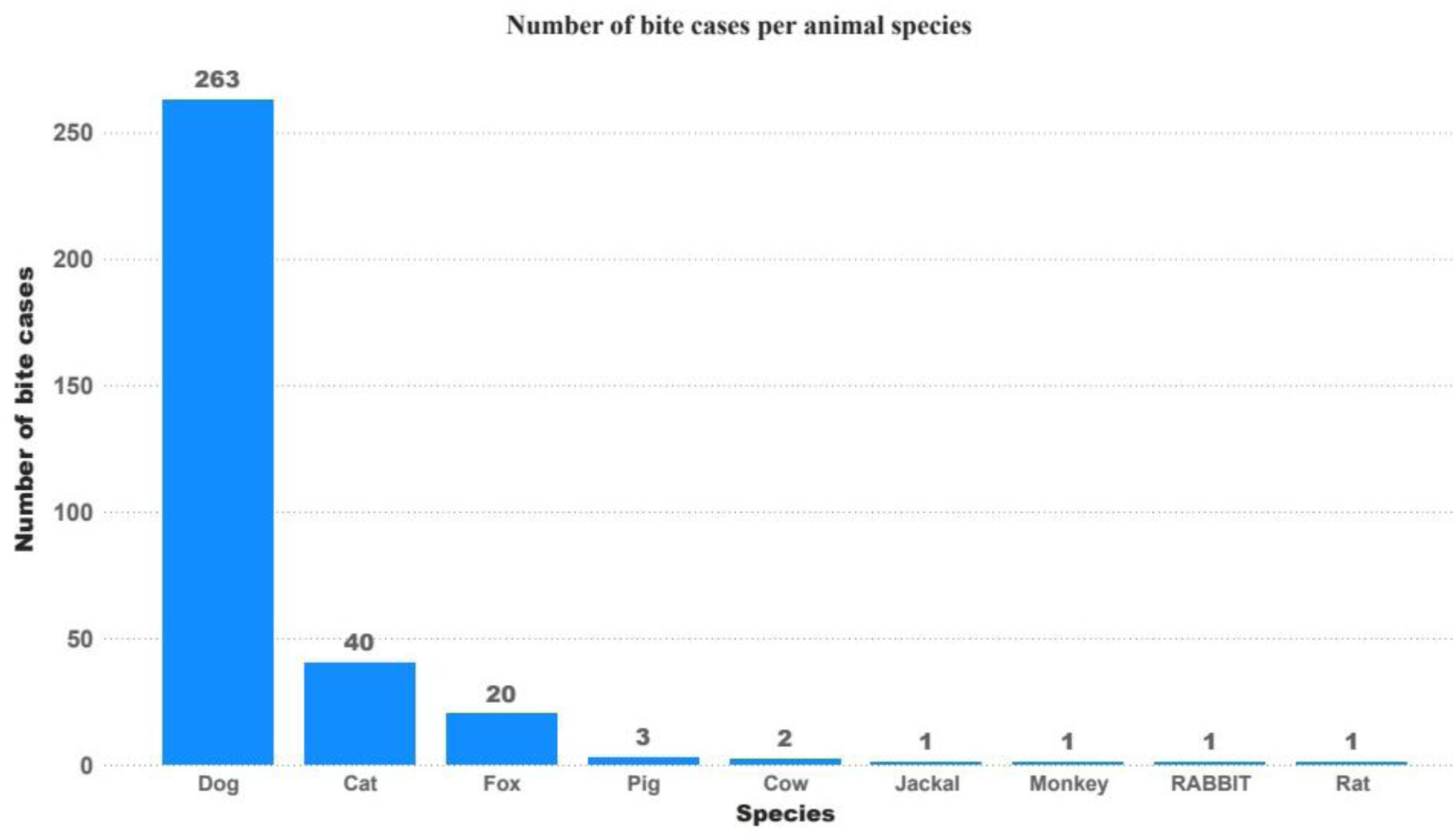
Number of bite cases per animal species

### Prevalence of suspected rabies exposure among animal bite human cases

The prevalence of suspected rabies exposure was 53.6% (95% CI: 46.8-60.3). The prevalences for the two years were not significantly different i.e. prevalence in 2023; 53.2% (95% CI: 43.8 – 62.3) and 2024; 53.9% (95% CI: 46.7-60.9).

Suspected rabies exposure cases appear to cluster around the main roads. Riverine areas highlight lower occurrence compared to non-riverine areas. Suspected rabies exposure cases occur across the full NDVI spectrum but rarely adjacent to the dense forest area. The distribution of the bite case locations is shown below in *Figure 3*

**Figure 3:**
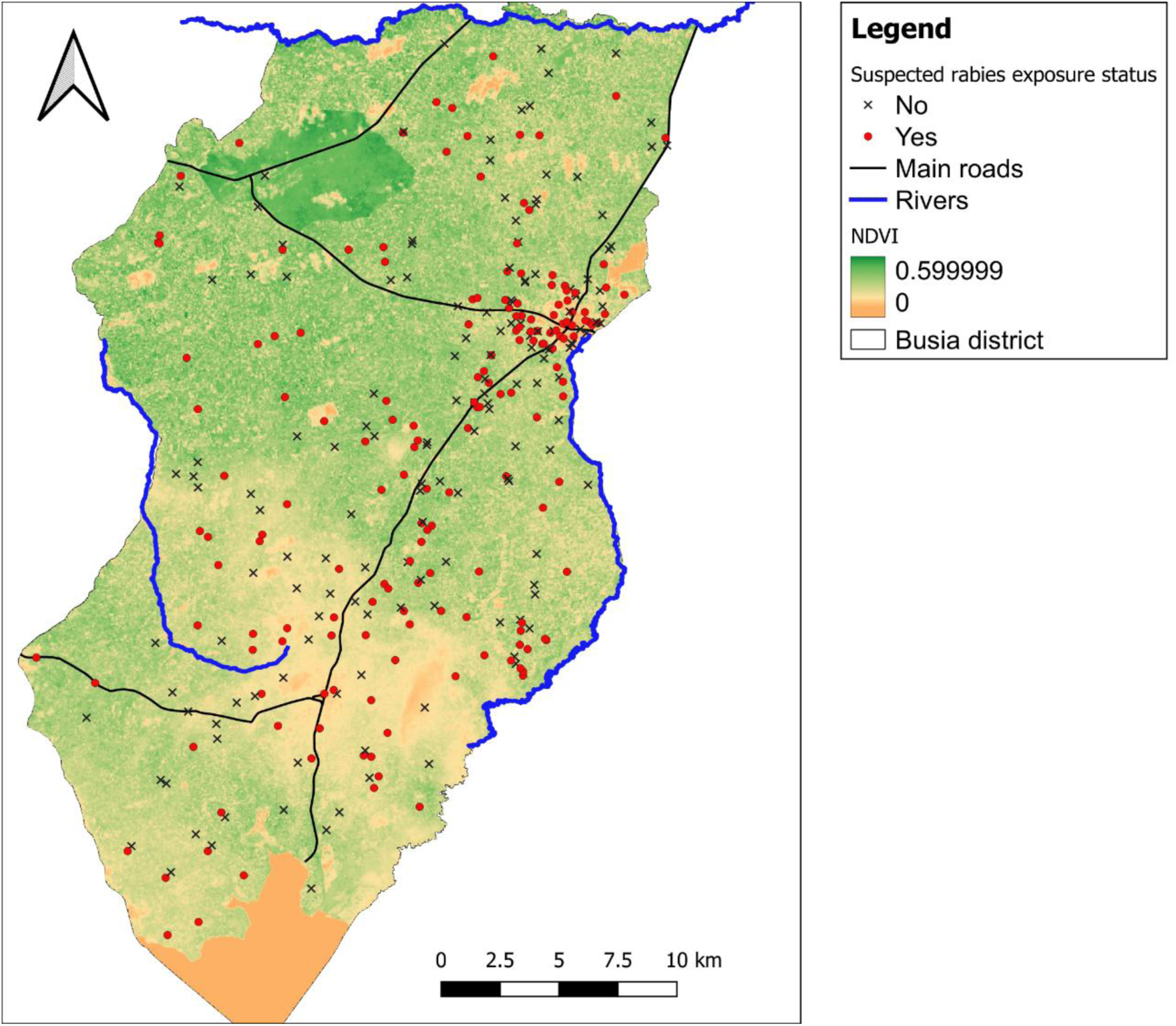
Map of Busia district showing the distribution of the 332 bite cases by location

## Factors associated with the prevalence of suspected rabies exposure among animal bite human cases in Busia district

### Bivariate analysis for the sociodemographic characteristics

The sociodemographic variables with a p-values less than 0.2 were: participants’ residence, household head education, family size, marital status of the household head and religion. The details of the variables are displayed in Table 3 below;

**Table 1:** Sociodemographic characteristics of the 332 bite cases in Busia district for the period 2023-2024;

| Variable | n (%) | Median (IQR) |
| --- | --- | --- |
| **Age of participant affected** |  | 18 (9-36) |
| **Age group household head** |  |  |
| 18 to 35 | 100 (30.1) |  |
| 36 to 59 | 185 (55.7) |  |
| 60 and above | 47 (14.2) |  |
| **Participant gender** |  |  |
| Male | 204 (61.4) |  |
| Female | 128 (38.6) |  |
| **Participant education** |  |  |
| No formal education | 53 (16.0) |  |
| Primary | 192 (57.8) |  |
| Secondary | 77 (23.2) |  |
| Tertiary (University/Diploma) | 10 (3.0) |  |
| **Household head education** |  |  |
| No formal education | 23 (6.9) |  |
| Primary | 40 (12.1) |  |
| Secondary | 132 (39.8) |  |
| Tertiary (University/Diploma) | 137 (41.3) |  |
| **Residence** |  |  |
| Rural | 295 (88.9) |  |
| Urban | 37 (11.1) |  |
| **Family size** |  | 6 (4-9) |
| **Marital status of household head** |  |  |
| Married | 290 (87.3) |  |
| Not married | 42 (12.7) |  |
| **Religion** |  |  |
| Christian | 309 (93.1) |  |
| Muslim | 22 (6.6) |  |
| Others* | 1 (0.3) |  |

\*Others; Traditional healer

**Table 2:**
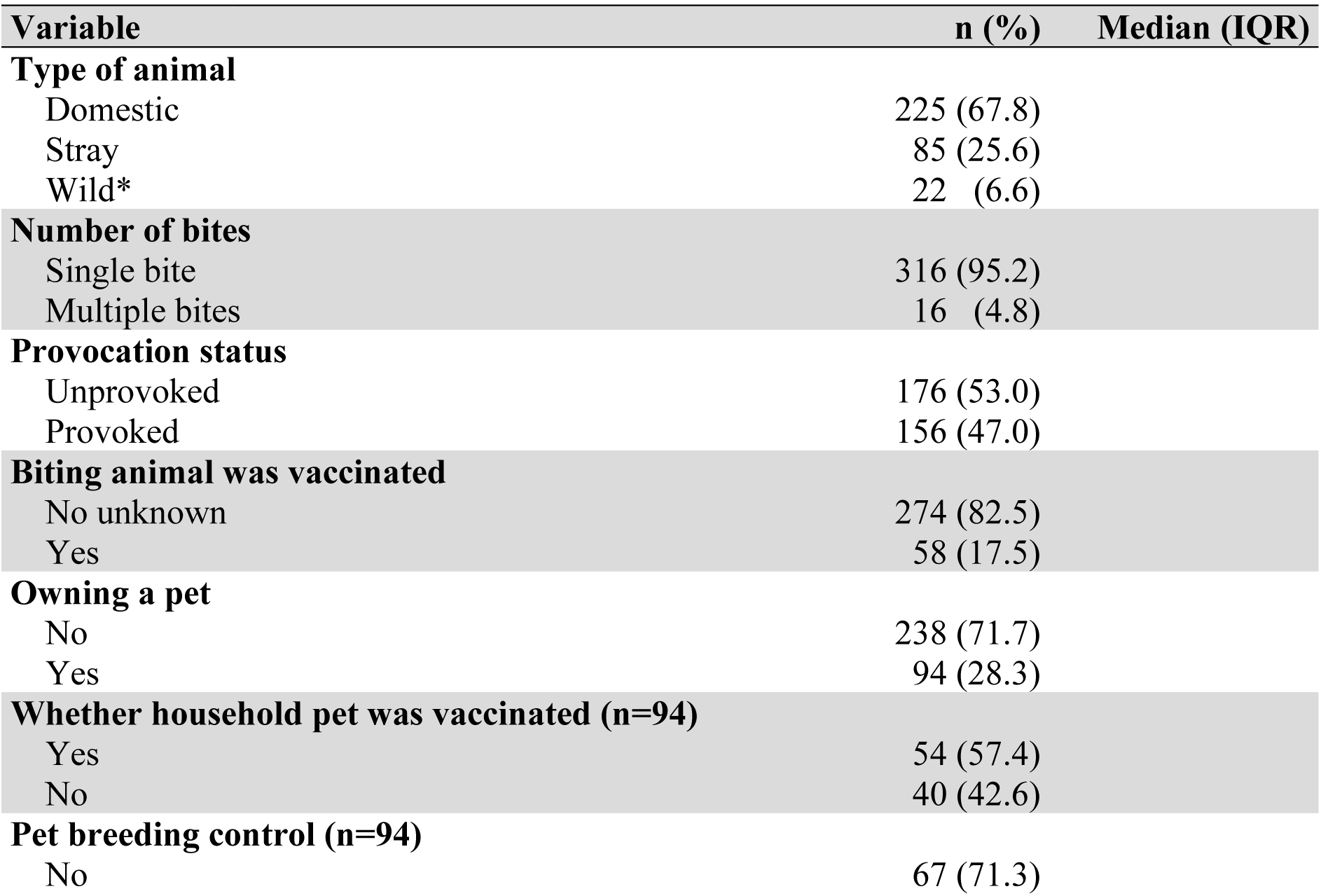
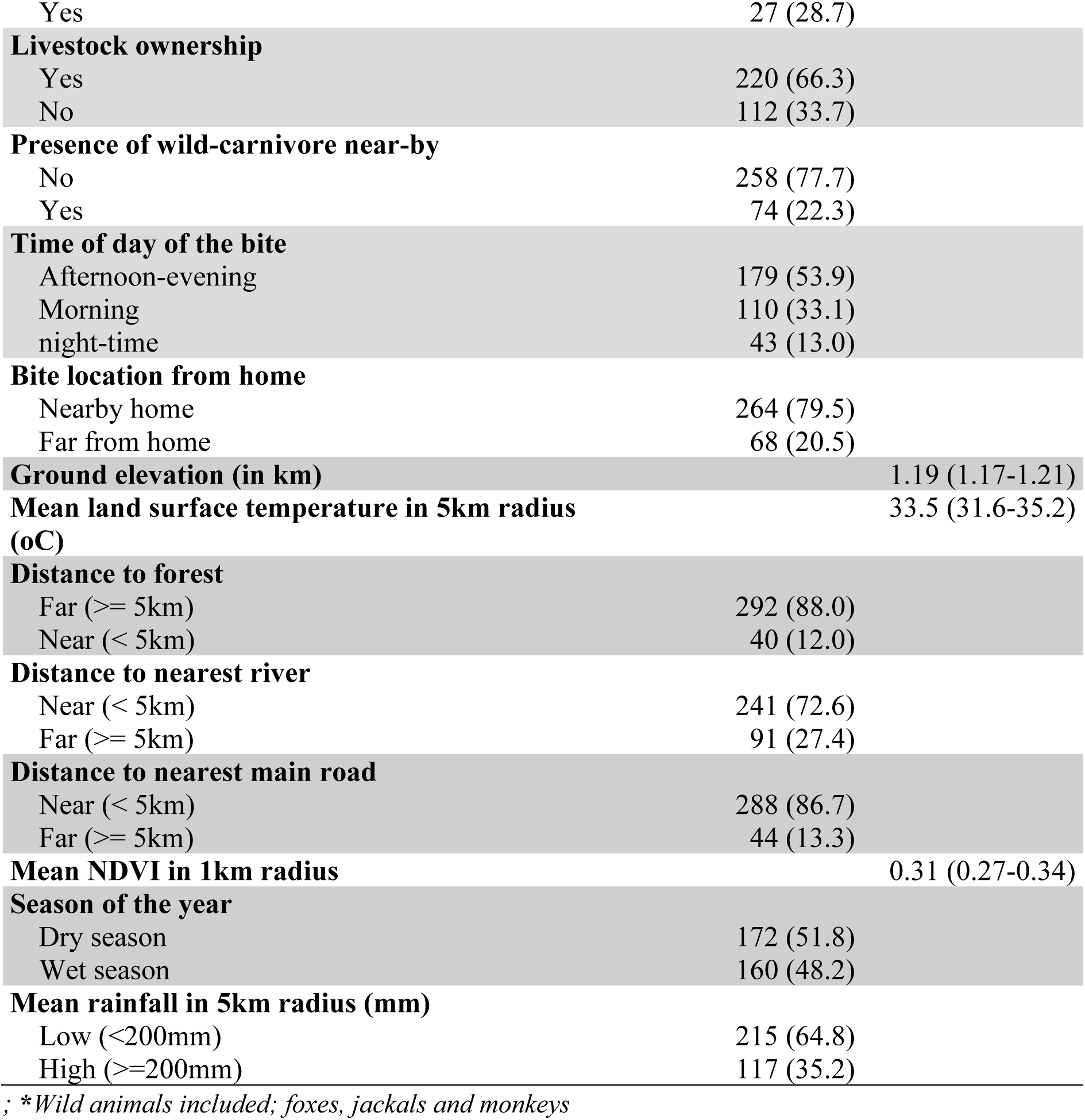
Animal-related and environmental characteristics for the 332 bite cases that occurred in Busia District; 2023-2024.

**Table 3:**
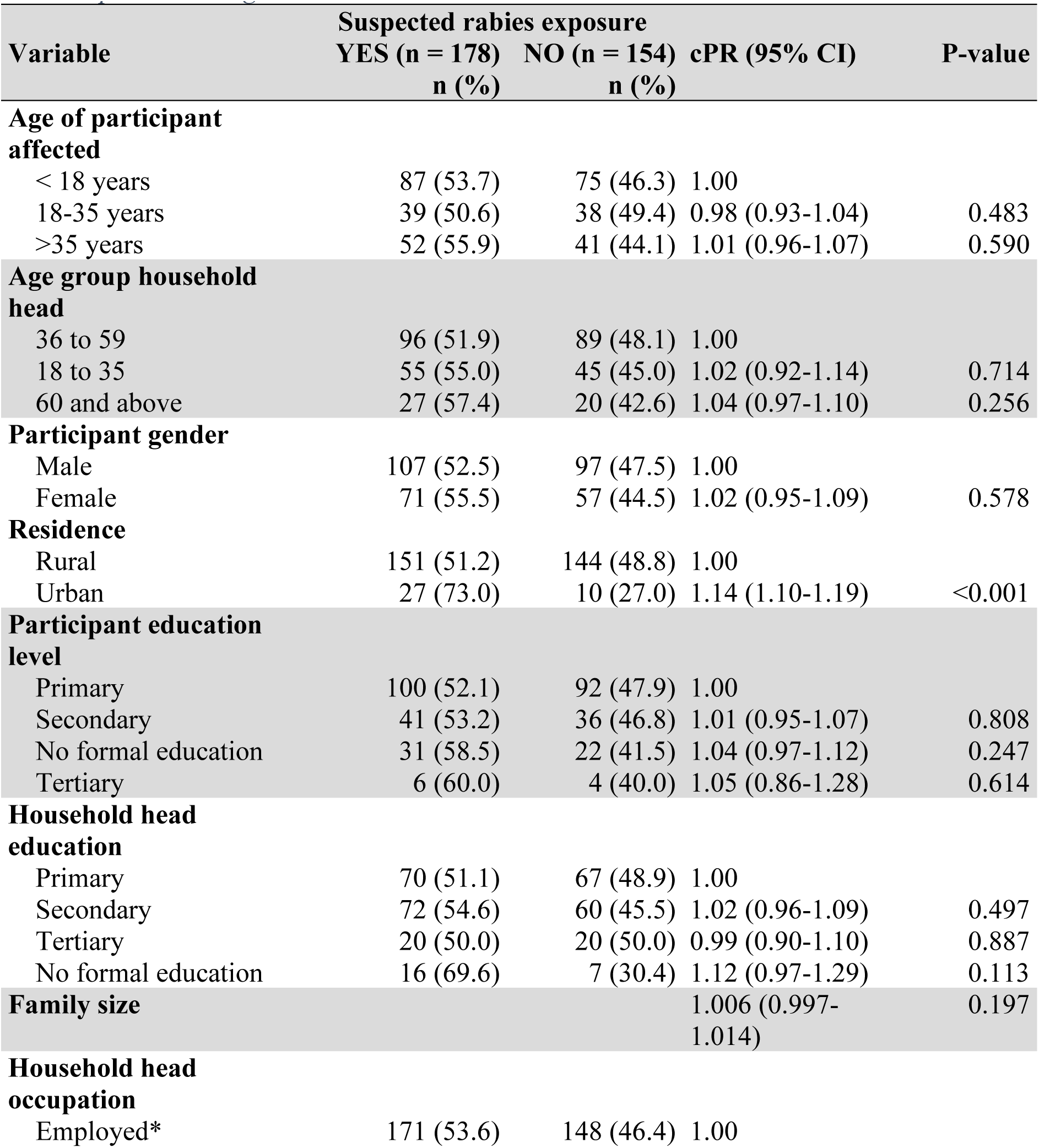
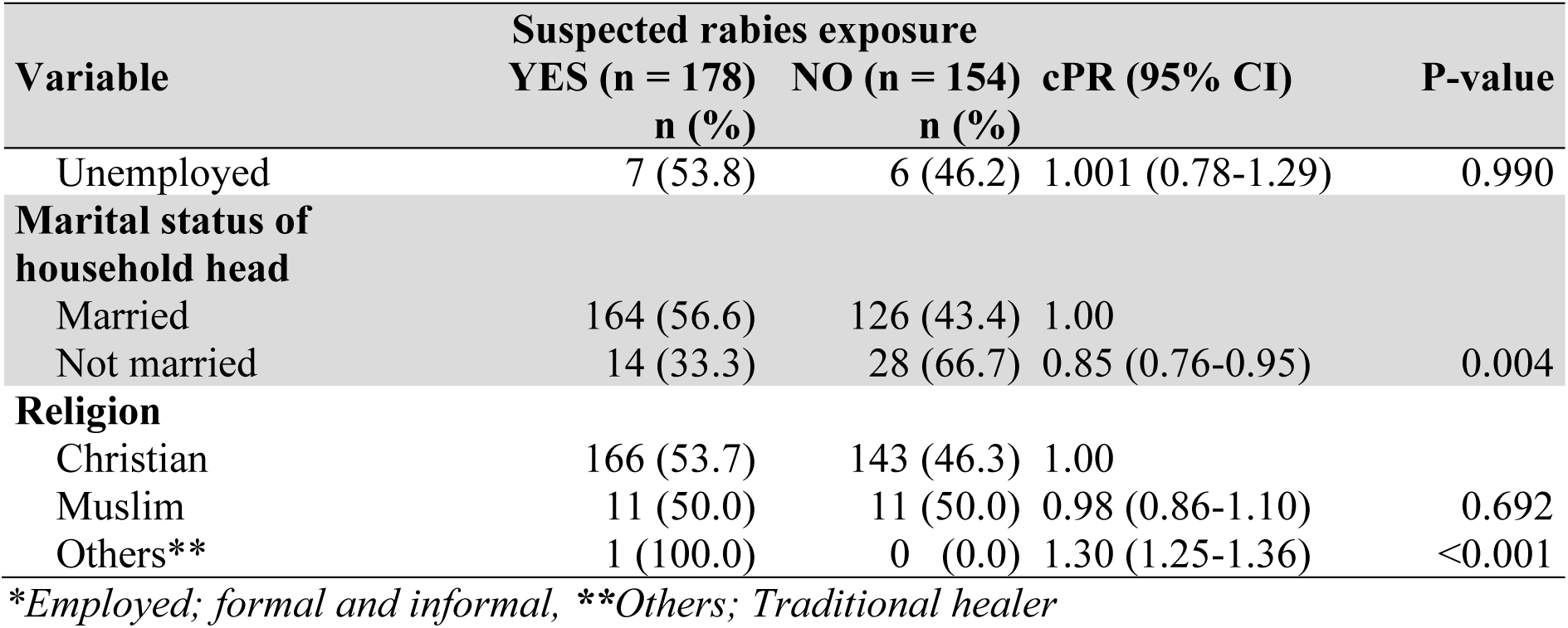
Bivariate analysis for the sociodemographic characteristics associated with suspected rabies exposure among the 332 animal-bite human cases in Busia district between 2023-2024.

### Bivariate analysis for animal-related and environmental characteristics

The variables that had p-values less than 0.2 were the type of animal, provocation status, vaccination status of the biting animal, owning a household pet, presence of a wild carnivore nearby, bite location from home. The environmental characteristics with p-values less than 0.2 were ground elevation, mean NDVI in 1km radius, distance to the nearest river and season of the year. The detailed results of the bivariate analysis for the animal and bite-related characteristics are shown in Table 4 below.

**Table 4:**
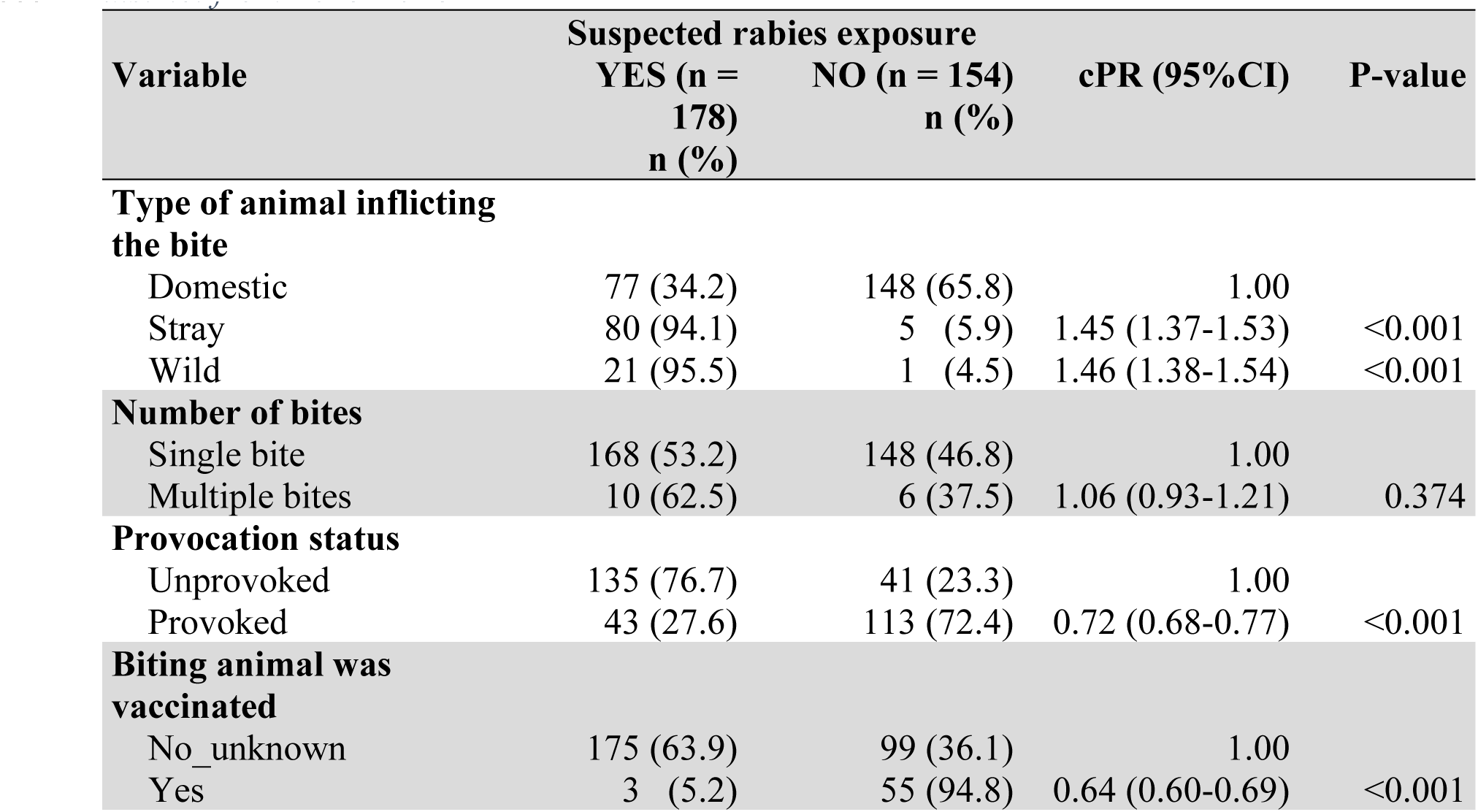
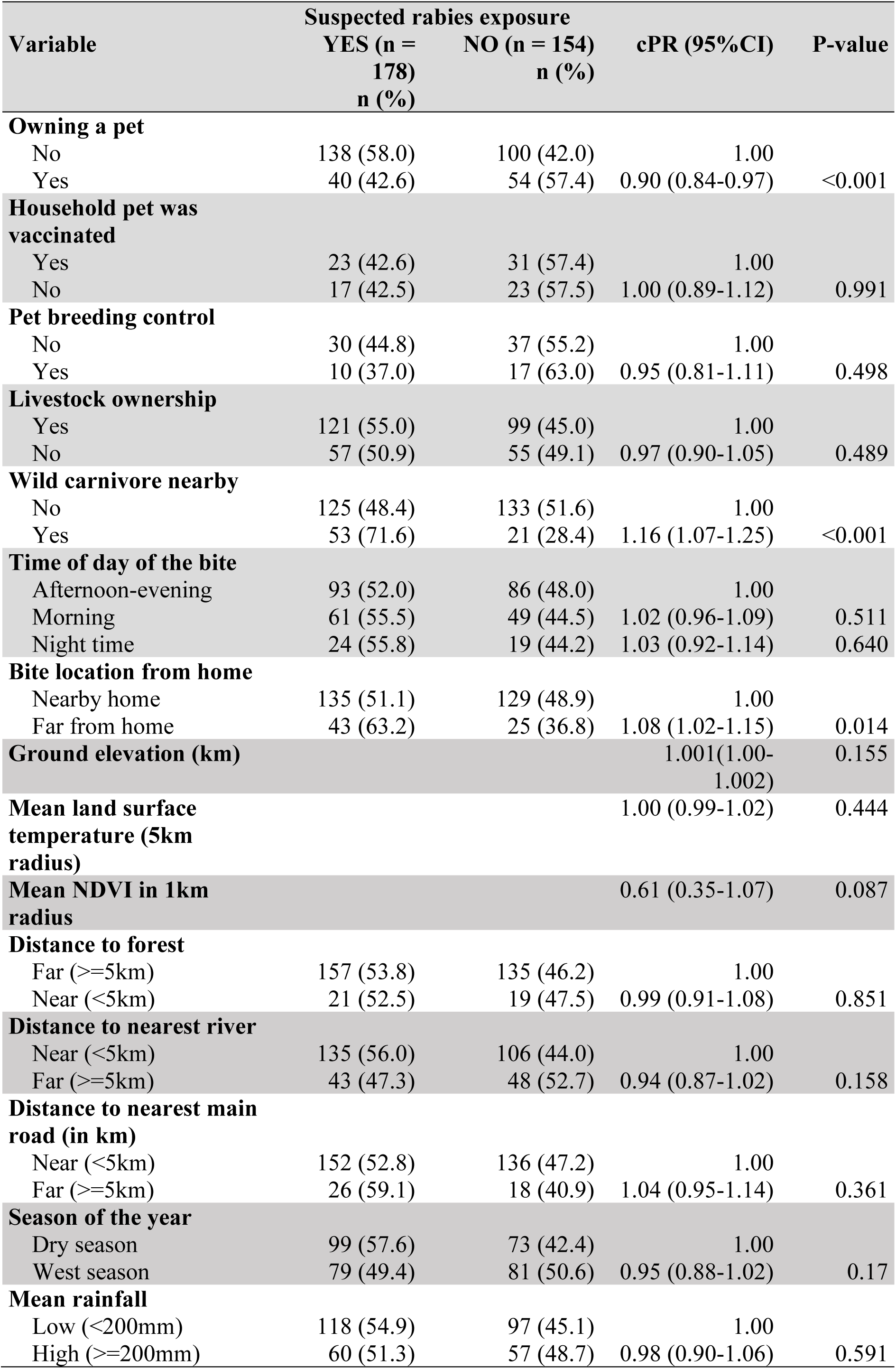
Bivariate analysis for the animal and bite-related and environmental characteristics associated with suspected rabies exposure among the 332 animal-bite human cases in Busia district from 2023-2024.

## Multivariable analysis results

Factors significantly associated with suspected rabies exposure included: residence, type of animal, provocation status, biting animal vaccination status, and distance to the nearest river. Table 5 below summarises the multivariable analysis results.

**Table 5:**
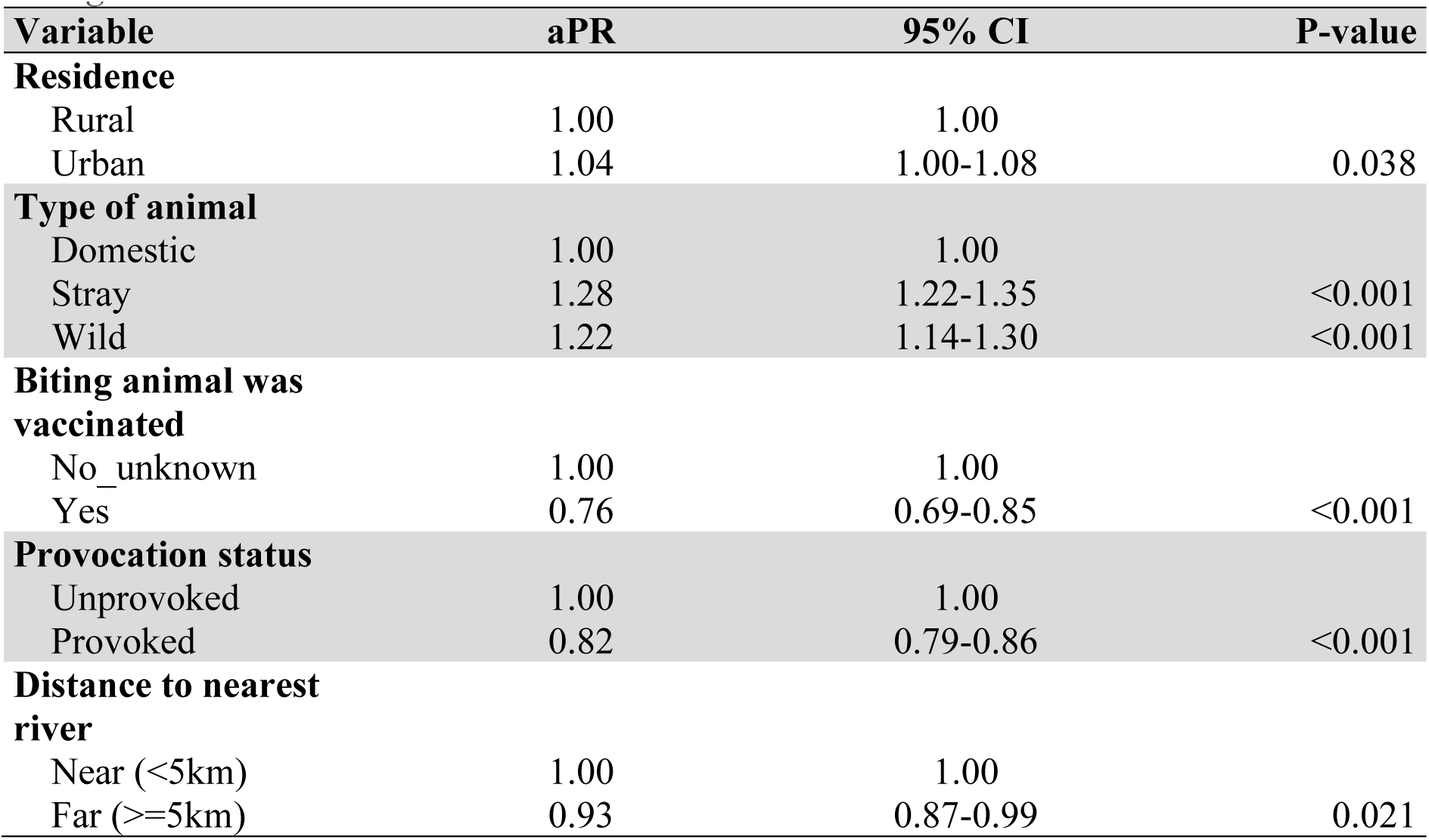
Multivariable analysis results for factors associated with suspected rabies exposure among 332 animal-bite human cases in Busia district: 2023-2024.

| Variable | aPR | 95% CI | P-value |
| --- | --- | --- | --- |
| **Residence** |  |  |  |
| Rural | 1.00 | 1.00 |  |
| Urban | 1.04 | 1.00-1.08 | 0.038 |
| **Type of animal** |  |  |  |
| Domestic | 1.00 | 1.00 |  |
| Stray | 1.28 | 1.22-1.35 | <0.001 |
| Wild | 1.22 | 1.14-1.30 | <0.001 |
| **Biting animal was vaccinated** |  |  |  |
| No_unknown | 1.00 | 1.00 |  |
| Yes | 0.76 | 0.69-0.85 | <0.001 |
| **Provocation status** |  |  |  |
| Unprovoked | 1.00 | 1.00 |  |
| Provoked | 0.82 | 0.79-0.86 | <0.001 |
| **Distance to nearest river** |  |  |  |
| Near (<5km) | 1.00 | 1.00 |  |
| Far (>=5km) | 0.93 | 0.87-0.99 | 0.021 |

## Prevalence of delayed post-exposure care-seeking among suspected rabies exposure bite cases

The prevalence was 23.0% (95% CI: 16.5-31.1). There was no statistically significant difference in the prevalences between years, although the prevalence was higher in 2023 (26.8%; 95% CI: 19.3-36.0) compared to 2024 (20.7%; 95%CI: 13.0-31.3).

The proportion of delayed post-exposure care-seeking was higher among those not suspected of rabies exposure i.e. 27.9% (95% CI: 19.5-38.3) compared to those suspected of rabies exposure, however, this difference is not statistically significant.

## Discussion

This study found a prevalence of suspected rabies exposure of 53.6% (95% CI: 46.8-60.3) associated with factors including residence, type of animal, provocation status, vaccination status of the biting animal and distance to the nearest river. Additionally, the prevalence of delayed post-exposure care-seeking was 23.0% (95% CI: 16.5-31.1).

The high prevalence of suspected rabies exposure observed implies that, on average, at least one out of every two bite incidents involved suspected rabies exposure. A possible explanation for this striking statistic is that Busia district was identified to be endemic with animal rabies (9), a situation that likely spurred public awareness campaigns and improved health-seeking behaviours following exposure. Furthermore, domestic-wild animal (foxes and jackals) interactions and vaccination rates below 70% further contribute to the high suspicion of rabies among the animals involved in these bites. These findings highlight a significant rabies risk in Busia district, posing a serious challenge in achieving the global goal of eliminating rabies deaths by 2030 especially in endemic rural areas.

In this study, individuals referred for PEP rabies vaccination were classified as having suspected rabies exposure. It was not explicitly clear whether WHO bite wound categorisation (1) was utilized as it wasn’t included on the PEP referral checklist. However, it is quite unlikely that category I exposures (i.e. contact of animal saliva on intact skin) would be reported to the DVO as they don’t lead to breakage of the skin. This implies that category II and III exposures (i.e. nibbling or single or multiple transdermal bites) were more likely to reported which aligns with WHO’s recommendation. Vaccination history, animal disposition and traceability are sometimes unclear especially in rural areas due to poor record keeping and unrestricted animal movements which are important during rabies risk assessment (23). This can lead to the possibility of either over- or under-referral of some of the bite exposures. This introduces differential misclassification of the suspected rabies exposure status hence the prevalence estimate is either over- or underestimated. However, careful clinical history about the animal involved is considered following bite exposure using the checklist to try to rule out cases not suspected of rabies which could minimize this bias.

This prevalence is higher compared to that of 10.47% identified by Azalu et al, 2024 which was carried out in North West Ethiopia (19). The high prevalence in this study could be attributed to the fact that bite cases reported at the district veterinary department were considered rather than those in the community. This was the case for the study in North west Ethiopia where community households were considered (19) leading to a significantly lower prevalence. Additionally, Busia district is also known to be endemic with animal rabies with reports of confirmed animal rabies cases in the past. This together with sensitization campaigns have improved awareness to rabies among the people which increases the likelihood of seeking care after bite exposure. Despite a similar definition of suspected rabies exposure, the study in Mukah division, East Malaysia, had a significantly lower prevalence (17.72%) possibly because the study was carried out following a suspected rabies case in a country that had been declared rabies free (20).

Among the bite cases, suspected rabies exposure was 4% more prevalent among urban residents compared to those in rural areas. This is contrary to what has been reported in previous literature for higher rabies risk to be common among rural settings compared to urban areas (24).

This difference could be attributed to the fact that the incidence of dog bites in urban settings in higher compared to rural settings due to higher dog populations and higher population density (25). This increases the frequency of bite incidents involving stray animals especially dogs and cats that usually lack a documented history of rabies vaccination. This results in bite cases being more likely classified as suspected rabies exposures. The urban area i.e. Busia municipal council is closer to the veterinary office and therefore has better access to health education services compared to rural areas. This would result in higher reporting rates by urban residents compared to those in rural areas which would slightly overestimate the measure of effect. Nevertheless, this finding implies that areas of high population density are more prone to suspected rabies exposure bites compared to those with low population density. Despite the need to increase the rabies vaccination rates among animals, special care should be placed in urban settings due to faster transmission per unit area.

The likelihood of suspected rabies exposure was 28% and 22% higher among bite cases for individuals affected by stray and wild animal bites compared to bites inflicted by domestic animals respectively. This finding is similar to the finding in the study carried out in Mukah division East Malaysia where it was identified that free-roaming and stray dogs posed a higher risk of rabies exposure compared to domestic pets. This finding is expected since stray animals are known to be unvaccinated and therefore are at high risk of developing clinical rabies (20). Additionally, wild carnivores are known to be reservoirs of rabies infection which later spread to the stray animals which freely roam within the communities. Stray and wild animals freely roam in search of food which leads them toward human settlements where bite incidents may occur as a result of protective or rabies associated aggression. This could also lead to infection of domestic animals in the rural communities perpetuating animal rabies transmission. Since these animals then run free following bite exposure, their actual rabies status is unclear leading to suspicions of rabies exposure (26). This dynamic is exacerbated in areas which are predominantly rural communities of where majority of the bite cases occurred. The finding indicates existing wild-domestic animal interactions which maintain unchecked rabies transmission among the animal population of Busia district. This finding is valid and aligns with global epidemiological patterns of rabies where unvaccinated animals, particularly stray animals and wildlife, are primary sources of human exposure. Therefore, there is a need to improve vaccination rates among domestic animal owners to improve immunity against the rabies virus if infected by the stray or wild life.

Suspected rabies exposure was 16% less likely among bite incidents that were provoked compared to those that were unprovoked. This finding is similar compared to the study carried out in Thailand where a bigger proportion of the category II and III exposures (suspected rabies exposure) were unprovoked although this was descriptively presented (27, 28). This is expected since unprovoked bites are part of the case definition of a clinical rabies case according to WHO (1). Although unprovoked bites are linked to rabies, such bites are commonly observed in the furious form of the disease. However, the paralytic form of rabies is the most common which may not be associated with unprovoked bites (29). This explains why some bite cases were suspected for rabies exposure even though provoked bites were involved. This shows that whether provoked or unprovoked, bite exposures pose a substantial risk for rabies exposure. The finding could be subject to reporting bias on the basis of the provocation status; however, unprovoked aggression would generally be clearly pointed out. Therefore, people of Busia district should be given comprehensive sensitization on the presentation of a rabid dog so that they exercise caution when handling animals with suspected rabies signs.

Suspected rabies exposure was 23% less likely among bite incidents that were inflicted by animals with a known history of vaccination. Rabies vaccination is known to be protective once done timely among domestic dogs and cats. Unvaccinated animals are highly susceptible to developing clinical rabies once infected and therefore individuals with bites inflicted by unvaccinated animals were suspected to be exposed to rabies (30). However, there have been reports of vaccination failure leading to animals exhibiting signs of rabies post-infection when vaccinated (28, 31). Additionally, it is also not clear whether the vaccination history provided for all bite cases was followed up with a certified vaccination card. This would mean that vaccination history of the biting animal would be subject to self-reporting bias at the point of risk assessment. The reasons highlighted could explain why some individuals bitten by animals that were reported to have been vaccinated were suspected to be exposed to rabies. Despite this, it is therefore advisable that most owned dogs are vaccinated annually against rabies as this improves protection against rabies exposure.

Bite cases that occurred far from the rivers i.e. above 5km were 7% less likely to be suspected of rabies exposure compared to those that occurred near the rivers. This is probably due to wild carnivore habitats towards the riverside areas especially for solitary wild carnivores like the foxes. This leads to increased suspected rabies occurrences among stray animals in the riverside areas due to wild-domestic carnivore interaction. This finding is supported by the study by Smith et al, 2002, who identified a decline in the effect of rabies occurrence with increasing distance from the river (32). This is contrary to what was reported by Yu *et al*, 2020 who highlighted increased rabies occurrence in areas far from major rivers. He added that rivers act as natural barriers to rabies transmission by restricting migration of these wild life (12). The geography of Busia district is unique with major rivers to its north, south east and western borders. This concentrates the bite incidents within the district however, with increased human settlement away from these riverine areas, solitary wild carnivores and stray animals could utilise them as habitats. Bites close to the riverside areas are then more likely to be associated with wild carnivore bites resulting in more suspected rabies exposure cases. Therefore, riverside areas in Busia are possible hotspot areas for rabies transmission. It is imperative that wild carnivore populations and migratory behaviour are investigated with the help of government wildlife authorities to help understand their dynamics in relation to rabies exposure.

The prevalence of delayed post-exposure care-seeking among those with suspected rabies exposure bites was 23.0% (95% CI: 16.5-31.1). This implies that at least 1 in every 5 suspected rabies exposure cases delayed to seek for post-exposure care to initiate post-exposure prophylaxis. This result is true since the dates of bite exposure and reporting for PEP referral are recorded for each bite case. However, it is lacking in precision due to relatively small sample size of suspected rabies exposures identified in this study. This finding is similar to that carried out in India who observed a prevalence of 25.14% among animal bite victims visiting a tertiary hospital in Pune, India (33). However, this prevalence is significantly lower than that reported in the study carried out in Arua district where about 69% delayed (22). This is probably due to previous reports highlighting more suspected rabies deaths in the Eastern region (34) and confirmed animal rabies in Busia (9) compared to other regions. This has encouraged annual rabies vaccination and awareness campaigns that have improved health seeking behaviour among residents. This enhances the recognition of the perceived susceptibility and severity following exposure as well as the associated benefits of seeking for PEP rabies vaccination. This also explains the similarity to the study in India because both regions of Busia district and India are known to be endemic with animal rabies. Despite the relatively low prevalence of delayed post-exposure care seeking, substantial delays in rabies PEP initiation could be observed if individuals do not initiate rabies PEP on the day of referral (35). This highlights existence of potential barriers to seeking PEP such as perceived costs and knowledge regarding timely administration of rabies PEP therapies. It could also highlight the use of alternative methods of therapy following bite exposure which could contribute to these delays. Therefore, there is a risk of rabies PEP failure even after initiation which has been linked with delays in PEP post-exposure seeking (1).

## Study limitations

Bite cases not reported at the DVO may differ in suspected rabies exposure status compared to those reported at the DVO. This could be in terms of perceived risk and differences in health seeking behavior among bite victims and their families. This selection bias is likely to be minimal since annual rabies vaccination and awareness campaigns are done in Busia district for example during the World rabies day. This results better bite incident reporting rates with a small proportion of bite cases not reporting to the DVO. The use of consecutive sampling introduced non-random selection of participants; however, the use of proportionate stratified sampling was used prior to record selection which ensured representative distribution of the bite cases.

Information bias i.e. in form of misclassification especially during risk assessment of the bite incidents. This could be as a result of variation in the risk assessment judgements by different veterinarians which could have introduced differential misclassification. Veterinarians involved in risk assessment receive comprehensive risk assessment trainings on rabies in order to improve reporting of this notifiable zoonotic disease. Additionally, self-reporting bias could be been introduced especially with regard to exposure variables which required the participant to recall issues around the bite incident. However, this is likely to be minimal since a time frame of two years prior was utilized. Traumatic events of which animal bites are included are less likely to be forgotten.

Although, the discussed biases could impact the internal validity of this study, their influence on the concluded measures is likely to be low. This limits the generalizability of these findings to predominantly rural areas with similar endemicity, weather patterns, and geographical features.

## Conclusion

The prevalence of suspected rabies exposure among animal-bite human cases in Busia district from 2023 to 2024 was high since for every 2 bite cases at least 1 would be suspected to be exposed to rabies. This high prevalence suggests that without prompt post-exposure care, roughly half of the bite victims could progress to clinical rabies. Suspected rabies exposure prevalence was associated with residence, the type of animal, provocation status, vaccination status, and distance to nearest rivers. These factors are multidimensional i.e. demographic, animal-related and environmental displaying a One health paradigm. The prevalence of delayed post-exposure care-seeking among those with suspected rabies exposure was relatively low i.e. for every 5 suspected rabies exposures, at least 1 delayed to seek post-exposure care. District Local Governments need to ensure availability of PEP rabies vaccines in areas of high rabies exposure prevalence. We recommend strengthening animal rabies vaccination campaigns to reduce infection transmission among domestic animals in the district. Trace, monitor and report wild carnivore movements to establish numbers and hot spot areas to advise on rabies exposure risk in Busia. The National One Health Platforms should strengthen and facilitate district one health collaborations to include human, animal and environmental variables as part of routine health surveillance of rabies-related cases. This can aid in monitoring trends as well as hot spot areas for establishment of targeted interventions. Further research can benefit from investigating potential barriers and facilitators with regard to PEP access in endemic regions.

## Ethical considerations

Ethical approval was obtained from School of Medicine Research and Ethics Committee (SOMREC) under protocol number: Mak-SOMREC 2024-1201. A waiver of informed consent was also sought from SOMREC as the study involved use of PEP referral records. Administrative clearance was sought for from the Busia District Chief Administrative Officer (CAO) and District Veterinary Office (DVO). Written informed consent and assent to participate in the study was sought from the animal bite cases.

## Consent for publication

Not applicable

## Availability of data and materials

The datasets used and/or analysed during the current study are available from the corresponding author on reasonable request.

## Author contributions

**D.W –** conceived the research idea, wrote the protocol, designed the study database and collection tools. He led the data collection, performed the data analysis and compiled the first draft of the research manuscript, **J.K** (**1**) **& H.K –** assisted in protocol, data collection tool design and field data collection procedures, **S.P.O** –assisted with collection of environmental variables and use of QGIS ,**H.U & D.M –** participated in the protocol design and statistical analysis planning, **A.K & C.K –** participated in proof reading the manuscript, **S.N** & **L.K.K –** assisted in the statistical analysis and drafting the manuscript, **I.N –** polished the research idea, supervised the design of the protocol and collection tools, supervised compilation of the manuscript, **J.K** (**2**) **–** refined the research idea, proof read the protocol, supervision of the study and review of the manuscript.

## Acknowledgements

We are grateful to the Busia District local government especially the Veterinary department for their assistance in field data collection. Special thanks to the lecturers and colleagues at the Department of Clinical Epidemiology and Biostatistics, College of Health Sciences, Makerere University for the skills and knowledge imparted.

## LIST OF ABBREVIATIONS

CHW: Community Health Worker
CHIRPS: Climate Hazards Centre InfraRed Precipitation with Station data
CNS: Central Nervous System
DVO: District Veterinary Office
GDP: Gross Domestic Product
GIS: Geographical Information System
GPS: Geographical Positioning System
HBM: Health Belief Model
HC: Health Centre
NDVI: Normalised Difference Vegetation Index
OPD: Outpatient Department
PEP: post-exposure prophylaxis
PI: Principal Investigator
PR: Prevalence Ratio
QGIS: Quantum GIS
SOMREC: School of Medicine Research Ethics Committee
UCG: Uganda Clinical Guidelines
URCS: Uganda Red Cross Society
US: United States
UWA: Uganda Wildlife Authority
VO: Veterinary Officer
WHO: World Health Organisation

